# A multi-trait approach improves polygenic risk scores for chronic back pain across population-based and clinically ascertained samples

**DOI:** 10.1101/2025.08.27.25334588

**Authors:** Rachael O. Osagie, Goodarz Koli Farhood, Marc Parisien, Amandeep Kaur, Hsuan Megan Tsao, Benjamin Kaufman, Justin Pelletier, Claude Bhérer, Audrey V. Grant, Carolina B. Meloto

## Abstract

Chronic back pain (CBP) is a complex, heritable condition, and a leading cause of global disability. Previous genome-wide (GW) CBP polygenic risk scores (PRS) derived from a large-scale cohort have shown low discrimination without clinical validation. To improve PRS performance and clinical relevance, we applied Multi-Trait Analysis of GWAS (MTAG) to summary statistics from five genetically correlated traits of European-ancestry individuals with UK Biobank (UKB) CBP as the primary trait, including dorsalgia and chronic musculoskeletal pain (N(effective)=492,717). For comparison, we also constructed a single-trait PRS using UK CBP-only GW data (N=234,013). PRS construction parameters were optimized in an independent large-scale cohort, the Canadian Longitudinal Study on Aging (CLSA) via five-fold cross-validation using LD clumping and p-value thresholding. With covariate adjustment, the MTAG-PRS achieved an AUC of 0.603 (AUC = 0.621; AUPRC = 0.346; R² = 0.051) that was slightly better than the UKB-only PRS (AUC = 0.604; AUPRC = 0.330; R² = 0.038). External validation in CBP cases and controls from another large-scale cohort CARTaGENE) confirmed the MTAG-PRS robustness (AUC = 0.638; AUPRC = 0.335; R² = 0.064). Validation in clinician-ascertained CBP cases (GENE-PAR study) contrasted against an independent subset of CARTaGENE controls improved the MTAG-PRS performance beyond the threshold for clinical utility (AUC = 0.785; AUPRC = 0.616; R² = 0.306). GENE-PAR CBP cases in the top decile PRS also displayed greater burden of CBP symptoms. These findings demonstrate that leveraging genetic pleiotropy, coupled with rigorous phenotyping, moved CBP PRS to clinical utility.

## Introduction

Chronic back pain (CBP) is a prevalent, complex, and heritable condition that imposes a substantial burden on individuals, healthcare systems, and society at large. Globally, it is a leading cause of disability with far-reaching socioeconomic consequences[16]. Twin and family studies estimate CBP heritability at 40% to 68%[5; 8; 43; 59]. Nevertheless, polygenic risk scores (PRS), which aggregate risk alleles weighted by their estimated effect sizes, currently explain only a modest fraction of this heritability[58; 73], consistent with CBP’s highly polygenic architecture and the additional contribution of non-genetic influences, such as environmental and psychosocial factors[22; 26; 27; 68; 69]. The first genome-wide CBP PRS[73], derived from a single-trait UK Biobank study, achieved modest discrimination (AUC ≈ 0.56) and was evaluated only in population-based samples, underscoring the need for clinical validation.

Genome-wide association studies (GWAS) have progressively identified risk loci associated with CBP, expanding from the initial discovery of three loci (*SOX5*, *CCDC26/GSDMC*, *DCC*) in ∼158,000 Europeans to over 100 genomic regions in a multi-ancestry meta-analysis of more than 550,000 individuals[8; 67; 69]. However, as with many complex traits, these loci individually explain only a small fraction of the phenotypic variance. Empirical observations across diverse complex traits suggest that individual SNPs typically account for less than 0.05% of a trait’s variance, underscoring a highly polygenic architecture where genetic risk is distributed across thousands of small-effect variants[10; 66]. The highly distributed nature of the genetic signal not only necessitates large sample sizes but also highlights the value of multi-trait genetic analysis approaches, which leverage genetic correlation with related traits to increase discovery power and improve risk prediction[3].

CBP shares substantial, genome-wide genetic correlations with anthropometric, musculoskeletal, and neuropsychiatric traits[7; 26; 79]. Multi-trait GWAS frameworks[30; 37; 42; 60; 74] leverage this overlap by jointly analysing correlated phenotypes. Multi-Trait Analysis of GWAS (MTAG) is a meta-analysis approach with a primary trait of interest and other secondary traits that boost power to detect association with the primary trait. MTAG models the genetic covariance between traits, increases the effective sample size, elevates sub-threshold associations to genome-wide significance, and returns trait-specific effect estimates suitable for PRS construction[48; 74]. Empirical applications of multi-trait GWAS methods to back pain phenotypes have reported additional loci and improvements in predictive accuracy[6; 7; 79]. However, translation of MTAG-derived CBP summary estimates into clinically informative PRS for physician-ascertained cohorts remains unexplored. Therefore, we sought to leverage traits genetically correlated to CBP using MTAG to derive a CBP PRS and assess its predictive accuracy and phenotype variance explained in a large population-based sample. Next, we evaluated the external validity and clinical relevance of the MTAG-derived PRS in an independent, clinician-ascertained sample, including comparisons of pain-related clinical features across genetic risk strata.

## MATERIALS AND METHODS

### Study Overview

We combined five GWAS summary statistics, i.e., CBP from the United Kingdom Biobank[28] (UKB-CBP), CBP from the Cohorts for Heart and Aging Research in Genomic Epidemiology[69] (CHARGE-UKB CBP), dorsalgia from the FinnGen study[44], dorsalgia from the Million Veteran Program[38], and chronic musculoskeletal pain (CMSKP) from the UKB (UKB CMSKP), and applied MTAG to boost CBP locus discovery and account for population overlap. We then used the MTAG-derived CBP summary statistics to build a PRS in the Canadian Longitudinal Study on Aging (CLSA) via five-fold cross-validation, selected the optimal clumping parameters based on the area under the receiver operating characteristic curve (AUC), and applied those parameters to score all CLSA participants. Performance was further evaluated first in CARTaGENE, a population-based case-control cohort, and then in a separate dataset composed of clinician-ascertained CBP cases (GENE-PAR) and an independent subset of CARTaGENE controls. Finally, within the clinician-ascertained sample (GENE-PAR), we compared LBP characteristics and comorbid symptoms burden between the top and bottom deciles of the PRS.

### Study Populations

This study leveraged genome-wide data from four consortia, yielding five discovery GWAS summary statistics that served as base data for PRS construction. All discovery GWAS analyses and subsequent PRS analyses were restricted to participants of European ancestry (**Supplementary Figure 1**). A summary of cohort characteristics, including sample sizes and case definitions, is provided in **Supplementary Table 1**.

We also describe the three target datasets used for PRS evaluation, including one internal and two external datasets.

### Discovery Dataset 1: The United Kingdom Biobank – Chronic Back Pain (UKB-CBP)

The UKB is a large-scale, multimodal population-based study that involves approximately half a million participants aged 40-69 years at recruitment (2006-2010). UKB genotyping, imputation, and quality control (QC) are described elsewhere[28]. We defined two case-control phenotypes using baseline, self-reported pain measures. Briefly, CBP cases (n = 70,643) were individuals who reported back pain that interfered with usual activities in the last month (Field 6159) and, in a conditional follow-up question, indicated that their back pain had lasted for three months or longer (Field 3571). Controls (n = 163,854) were participants who reported no pain at any body site. Individuals who reported back pain lasting less than 3 months or selected “prefer not to answer” were excluded.

### Discovery Dataset 2: The United Kingdom Biobank – Chronic Musculoskeletal Pain (UKB-CMSKP)

In the UKB, CMSKP cases (n = 156,235) were defined as chronic pain (≥3 months) in at least one of four sites (back, knee, hip, neck/shoulder). Controls (n = 154,045) reported no pain at any site. UKB genotyping, imputation, and quality control (QC) are described elsewhere[28].

### Discovery Dataset 3: The Cohorts for Heart and Aging Research in Genomic Epidemiology - Chronic Back Pain (CHARGE - UKB)

The CHARGE - UKB meta-analysis pooled data from 16 population-based cohorts (15 CHARGE cohorts and one from the UKB interim release; n = 29,531 cases, n = 128,494 controls) and harmonized CBP definition across cohorts as follows: (1) ≥3 months of back pain, (2) ≥6 months of back pain, or (3) ≥1 month of back pain per year in consecutive years (≈ 12 months total). Controls were those with no back pain or pain of insufficient duration[69].

### Discovery Dataset 4: FinnGen Study

The FinnGen project (https://www.finngen.fi/en) integrates genotype data with national longitudinal health registry data for over 500,000 Finnish individuals. Phenotype definitions in FinnGen were defined using the Finnish version of the International Classification of Diseases (ICD-10) and harmonized with ICD-8 and ICD-9 where applicable to incorporate historical health records[44]. Dorsalgia cases were defined using the curated FinnGen endpoint M13_DORSALGIA, which corresponds to ICD-10 code M54 and encompasses a range of back pain diagnoses, including low back pain, sciatica, radiculopathy, cervicalgia, thoracic spine pain, and unspecified dorsalgia. We utilized GWAS summary statistics from Data Freeze 12, comprising 83,888 cases of dorsalgia and 353,224 controls. Controls were defined as individuals without any diagnosis under the broader M13_DORSOPATHY category. Genotyping, imputation, and quality control procedures have been described previously[44].

### Discovery Dataset 5: The Veterans Affairs (VA) Million Veteran Program (MVP)

We used GWAS summary statistics for dorsalgia in MVP. These data were obtained from a publicly available meta-analysis conducted by FinnGen, which combined results across MVP, FinnGen, and UKB cohorts (available at https://mvp-ukbb.finngen.fi). Specifically, we extracted the summary statistics corresponding to the MVP cohort by selecting the columns labeled “MVP_EUR” within the meta-analysis results. This subset included 221,759 dorsalgia cases and 187,718 controls. Phenotype harmonization across cohorts was conducted by the FinnGen consortium. Endpoints from UKB were included only if they exactly matched FinnGen definitions, ensuring strict alignment. FinnGen permitted inclusion of imperfectly matching endpoints if sample overlap exceeded 90%, while MVP endpoints were included regardless of matching status. Concordance of effect sizes at top loci was evaluated across cohorts to support phenotype alignment; however, no formal heterogeneity statistics or exclusion thresholds based on effect size discordance were reported. Genotyping, imputation, and QC procedures in MVP have been described elsewhere[38].

### Target Dataset 1: The Canadian Longitudinal Study on Aging (CLSA)

The CLSA is a multimodal, prospective study that enrolled ∼50,000 subjects aged 45-85 years, primarily of European ancestry (93%). Details on CLSA genotyping, imputation, and QC are described elsewhere[24; 64]. At baseline, the comprehensive CLSA cohort comprised 30,097 participants with complete questionnaire data. Of these, 5,109 participants reported back pain on most days for at least one month; indicated that the pain had persisted for three months or longer (duration ≥ 0.25 years); and confirmed that the pain had occurred within the previous year. Pain-free controls (n = 13,577) reported no history of back pain of this duration and stated that they were usually free of pain or discomfort. After merging the genetic and phenotypic data, we retained 3,201 participants with CBP and 9,484 control participants of European ancestry for the analysis.

### Target Dataset 2: CARTaGENE

CARTaGENE is a prospective, population-based cohort designed to investigate the link between genetic factors and chronic diseases in Quebec men and women aged 40-69 at recruitment[4]. Participants completed questionnaires, underwent physical measurements, and provided biological samples. Sociodemographic and lifestyle questionnaires were self-administered, and a trained interviewer administered the health questionnaire. Genotyping and QC information are described elsewhere[4; 41]. We used data collected about pain to identify individuals with CBP and pain-free controls. CBP cases (n = 632) were defined as participants who answered “yes” to the question: “Has a doctor ever told you that you have chronic back pain?” within the musculoskeletal/connective-tissue domain. Pain-free controls (n = 3,339) were participants who (1) reported being “usually free of pain or discomfort”, and (2) answered “no” to any doctor-diagnosed condition in the musculoskeletal/connective-tissue list. This dataset was used in two independent validation contrasts to calculate PRS performance metrics. The CARTaGENE - controls subset comprised pain-free individuals only that were used as controls for GENE-PAR cases, given that the latter is a case-only cohort and both populations are from the same geolocation and share similar genetic architecture (**Supplementary Figure 2**). After removing related individuals, we randomly sampled 864 CARTaGENE pain-free controls to compose this subset, achieving a 1:3 case-control ratio[40] relative to the 288 GENE-PAR chronic low back pain cases (n in the GENE-PAR + CARTaGENE – controls = 1,152). The CARTaGENE – case-control subset was comprised of cases and controls from CARTaGENE only. From the remaining 2,475 pain-free controls, we randomly sampled 2,319 to be contrasted against the 632 CARTaGENE cases to achieve a similar 1:3 case-control ratio (n in the CARTaGENE – case-control subset = 2,951) and ensure comparable PRS metrics. Participants with “don’t know” or “prefer not to answer” responses to any relevant item were excluded.

### Target Dataset 3: The GENEtic Variants Associated with PAin Reduction (GENE-PAR) Study

The GENE-PAR Study comprises a subset of individuals with clinician-ascertained chronic low back pain (CLBP), i.e., with a clinical diagnosis of CLBP given by a pain specialist, who participated in the Quebec Pain Registry (QPR, www.quebecpainregistry.com)[15]. Briefly, our team contacted QPR CLBP participants via mail and collected data and samples from them between 2015-2016. Participation in the GENE-PAR Study entailed donating a saliva sample, completing a questionnaire (the Canadian adaptation of the NIH Minimum Dataset for LBP research[20; 45]), and returning both to our team using a pre-paid envelope included with the mail. In total, we recruited 349 QPR CLBP participants, of which 327 provided completed questionnaires. We confirmed their CLBP status as per the NIH classification criteria using the first two questions of the Canadian adaptation of the NIH Minimum Dataset for LBP research (i.e., LBP that has been an ongoing problem for at least 3 months and has resulted in a problem on at least half of the days in the past 6 months)[20; 21]. Additional data collected with the Minimum Dataset included: average pain intensity in the past 7 days (0-10 numerical rating scale), the presence or absence of pain that spreads down the leg (sciatica pain), the presence or absence of one or more bothersome painful comorbidities (stomach pain, pain in arms, legs, or joints, headaches, and widespread pain; being bothered a little or a lot by one of these conditions characterized the presence of bothersome painful comorbidity), history of LBP surgery (i.e., having ever had a LBP surgery), pain interference (the PROMIS Short-Form-4a pain interference score)[1], ongoing treatments for LBP (opioids, infiltrations/injections, exercise therapy, or psychological counselling), LBP-related work absenteeism (yes to being off work or unemployed for 1 month or more due to LBP), LBP-related workers’ compensation benefits (yes to receiving or having applied for disability or workers’ compensation benefits because of LBP), physical function (the PROMIS Short-Form-4a physical function score)[1], depression (the PROMIS Short-Form-4a emotional distress or depression score)[1], sleep disturbance (the PROMIS Short-Form-4a sleep disturbance score)[1], kinesiophobia (agreeing to “it is not really safe for a person with my low back problem to be physically active”)[12; 34], pain catastrophizing (agreeing to “I feel that my low back pain is terrible and it’s never going to get any better”)[12; 34], history of LBP-related lawsuits and legal claims (being involved in a lawsuit or legal claim related to LBP), substance abuse (answering sometimes or often to “have you consumed alcohol or used drugs more than you meant to?” or to “have you felt you wanted or needed to cut down on your drinking or drug abuse?”). Next, the minimum dataset includes questions on sociodemographic data (i.e., age, sex, race, educational attainment, and employment status), as well as smoking status, and height and weight that were used to derive the body mass index (BMI). Neuropathic pain features were screened with the Douleur Neuropathique-4 (DN-4) questionnaire[9], and somatic symptom burden was quantified using the Somatization subscale of the Symptom Checklist-90 (SCL-90)[19]. These measures were used to characterize this clinical group of individuals with CLBP and to evaluate how their PRS scores relate to CLBP symptoms and comorbid symptoms.

DNA was extracted by the Genome Quebec Innovation Centre using QIAsymphony. Genotyping will be done by the Genome Quebec Innovation Centre, using the Axiom® Precision Medicine Research Array (Thermo Fisher Scientific). Arrays will be spiked with technical replicates to ascertain the integrity of genotyping plates. Following genetic QC, 288 unrelated participants remained for PRS construction.

### Target datasets 2 and 3: Genotype Imputation

To combine GENE-PAR cases with CARTaGENE controls for PRS construction, we imputed GENE-PAR and CARTaGENE genetic data (*de novo*) as described below.

Prior to imputation, chip controls from the GENE-PAR genotype dataset were removed, along with individuals of ambiguous genetic sex, using PLINK v1.96[63]. The CARTaGENE[4] genotype dataset was arbitrarily divided into two halves (N1 = 15,500, N2 = 13,056) to accommodate the Michigan Imputation Server’s limit on the number of samples per job[18]. Variants with a minor-allele frequency under 1%, a variant call rate below 95%, and a significant departure from Hardy-Weinberg Equilibrium (HWE) (p < 10^-25^) were filtered out[72]. Samples with a call rate under 95% were excluded from the analysis. Strand alignment and normalization were performed using bcftools, and chromosome names were reformatted to meet the requirements of the Michigan Imputation Server [17; 18].

Genotype imputation was performed on three datasets (the GENE-PAR cases and the two CARTaGENE batches) using the Michigan Imputation Server using the TopMed reference panel (TopMed-r-3)[18; 71]. The Michigan Imputation Server employs a hybrid approach, utilizing Eagle v2 for pre-imputation haplotype phasing and Minimac2 for genotype imputation [29; 47].

In each of the three imputed datasets, variants with imputation quality scores (R²) below 0.6 were excluded. The three file sets were then merged using bcftools, resulting in 19,618,805 shared variants. Principal component analysis was performed on a pruned version of the merged GENE-PAR + CARTaGENE dataset (LD correlation R² > 0.5, window size 50 kb, step size 5 kb; MAF>5%; SNP missingness > 1%; HWE p < 10^-25^) using PLINK v1.96 and FlashPCA v2[2; 63]. To identify and exclude batch-effected variants in the new merged dataset, a pairwise genome-wide association study was run between samples originating from GENEPAR and those from CARTaGENE[72]. A logistic regression was run with the first 20 principal components as covariates using PLINK v1.96[63; 72]. The logistic regression scheme to test for batch-effected variants in the merged imputed dataset is as shown below:

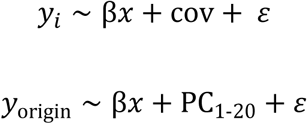

### Computational and statistical analysis

#### GWAS summary statistics

We assembled summary statistics for five European-ancestry GWAS (UKB CBP & CMSKP; CHARGE-UKB CBP; FinnGen dorsalgia; MVP dorsalgia). We performed UKB CBP and CMSKP scans using REGENIE v3.2.2[52], fitting linear mixed-effects models of SNP dosage on phenotype and adjusting for age, age squared, sex, genotyping batch, and PCs 1-40. CHARGE-UKB CBP, FinnGen, and MVP dorsalgia summary statistics were accessed from https://gwasarchive.org, https://r12.finngen.fi, and https://mvp-ukbb.finngen.fi, respectively.

#### SNP-based heritability and Genetic correlations with related traits

We applied linkage disequilibrium score regression (LDSC; v1.0.1)[13] to estimate the genetic correlation between UKB CBP, CHARGE-UKB CBP, FinnGen dorsalgia, MVP dorsalgia, and UKB CMSKP. LDSC estimates genetic correlation by exploiting the relationship between GWAS test statistics and linkage disequilibrium (LD), whereby a SNP’s association statistic reflects both its own causal effect and the effects of variants in LD with it. To ensure consistency across datasets, the effect alleles and non-effect alleles from the GWAS datasets were harmonized using the munge_sumstats.py script. We then used the ldsc.py script to compute SNP heritability and genetic correlations, referencing the pre-computed LD scores from the 1000 Genomes European data[32]. The detailed analytical procedure can be found at https://github.com/bulik/ldsc/wiki/Heritability-and-Genetic-Correlation.

#### Multi-trait analysis

To identify the optimal combination of genetically correlated traits for inclusion in our Multi-Trait Analysis of GWAS (MTAG), we systematically evaluated various trait groupings. Each combination was assessed based on predefined criteria, including improvements in the MTAG mean chi-square statistic, GWAS-equivalent sample size, and enhancements in polygenic risk score (PRS) performance across multiple clumping parameters. Based on these evaluations, we selected a five-trait model comprising UKB baseline CBP, CHARGE-UKB CBP, FinnGen dorsalgia, MVP dorsalgia, and UKB CMSKP for subsequent analyses.

MTAG jointly analyzes GWAS summary statistics from genetically correlated traits while accounting for sample overlap using bivariate linkage disequilibrium score regression (LDSC)[13; 74]. This approach provides trait-specific SNP effect estimates and pairwise genetic correlations among the input traits. We first harmonized all GWAS summary statistics to the same genomic coordinate system using the command line tool liftOver (https://genome-store.ucsc.edu/). The final MTAG analysis was restricted to the intersection of variants shared across all five datasets.

To evaluate the credibility of MTAG results, we assessed the maximum false discovery rate (MaxFDR) for each trait. MaxFDR reflects the worst-case expected proportion of false discoveries among genome-wide significant findings under the MTAG model[74].

In addition, following a hold-out approach used in prior large-scale GWAS[46] we conducted two replication analyses to evaluate the robustness and added value of the MTAG results. In the first analysis, we utilized genome-wide significant SNPs (p < 5 × 10⁻⁸) from the MTAG output for the UKB CBP trait, employing a four-trait MTAG model that excluded MVP dorsalgia to ensure sample independence. In the second analysis, we focused on genome-wide significant SNPs from the MTAG-enhanced CHARGE-UKB CBP results, selected because CHARGE-UKB was the lowest-powered input GWAS in the five-trait model. For both analyses, SNPs were matched by variant ID to the MVP dorsalgia GWAS summary statistics. A SNP was considered replicated if it showed a consistent direction of effect and nominal significance (p < 0.05) in the MVP dataset.

#### Polygenic Risk Score Development

We constructed polygenic risk scores (PRS) using the clumping and thresholding (C+T) approach[39] as implemented in PRSice-2[14]. After aligning alleles and filtering to autosomal SNPs with MAF ≥ 0.05, a total of 3,757,833 variants were retained for PRS construction.

We performed 5-fold cross-validation over 45 PRS models defined by all combinations of LD clump r² thresholds (0.1, 0.5, and 0.7) and window sizes (w_c_) (250 kb, 500 kb, 1000 kb) as hyperparameters[23; 62]. Within each fold and clumping configuration, PRSice-2 simultaneously calculated scores at eight predefined p value thresholds specified with --bar-levels (P_T_ = 1, 0.5, 0.4, 0.3, 0.2, 0.1, 0.05, 0.001); the P_T_ maximizing mean AUC across folds was selected. All PRS were standardized to Z-scores (mean = 0, SD = 1) within each fold. We then recalculated for the full sample using the optimal r², w_c,_ and P_T_ settings.

We compared mean age between cases and controls in CLSA and GENE-PAR + CARTaGENE controls using Welch’s two-sample unequal variances t-tests to determine whether age bias by phenotype warranted inclusion of a quadratic age term in downstream models.

To assess the association between PRS and CBP, we applied logistic regression models in the CLSA cohort, adjusting for age, age squared, sex, and the first 10 principal components (PCs) of ancestry. The model was specified as:

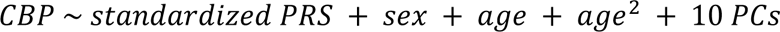

To allow for comparison with the recalculated PRS based on the MTAG boosted summary statistics, we derived a second PRS in the CLSA from a single-trait UKB CBP GWAS. Incremental predictive utility was assessed via Nagelkerke’s pseudo-R²: (i) covariates only; (ii) covariates + UKB PRS; (iii) covariates + MTAG PRS. The MTAG gain was defined as the difference between (iii) and (ii), with standard errors estimated by 1000-replicate bootstrap.

Discrimination was assessed using AUC by pROC[65] and area under the precision-recall curve (AUPRC) by PRROC[33]. AUCs were compared by DeLong’s test and calibration was examined using Brier scores[11]. To further evaluate predictive performance, we stratified participants into PRS quantiles and estimated odds ratios for each quantile compared to the middle 40-60% group. We additionally assessed case enrichment in the top 5% of the PRS distribution. Predictive performance was evaluated using standardized metrics[76].

#### External PRS validation

We first assessed the MTAG-derived PRS in the CARTaGENE – case-control subset (n = 2,951; 632 cases) and subsequently in the GENE-PAR + CARTaGENE – controls dataset (n = 1,152; 288 cases). Individual scores were calculated in PRSice-2[14] using the optimized parameters (r² = 0.5; w_c_ = 1000 kb; P_T_ < 0.1) and standardized to Z-scores for each analysis. We compared standardized PRS between cases and controls using Welch’s two-sample unequal variances t-test and reported the mean difference with 95% CI.

To compare genetic risk between groups, we contrasted PRS distributions in cases versus controls via Welch’s two-sample t-test, reporting mean differences with 95% confidence intervals. We then fitted three logistic regression models for CBP status: (1) PRS only (per 1-SD increase); (2) demographics-adjusted (PRS, age, age², sex); and (3) fully adjusted (demographics + PCs 1-10). For each model, we report odds ratios per 1-SD PRS increment, Nagelkerke’s pseudo-R² to quantify variance explained and discrimination metrics (AUC by pROC[65]; AUPRC by PRROC[33]). Brier scores[11] were estimated for all PRS models.

We next stratified participants into PRS deciles and tested enrichment of CBP cases in the top versus bottom decile using Fisher’s exact test, deriving decile-specific odds ratios from the resulting 2×2 contingency tables.

Finally, we compared clinical and psychosocial characteristics between the bottom and top PRS deciles among GENE-PAR cases [25; 56; 75]. Continuous measures were compared by Welch’s unequal variances t-tests, and categorical measures by Fisher’s exact tests.

## Results

### GWAS summary statistics

The UKB CBP GWAS identified 52 independent significant SNPs (P < 5 × 10⁻⁸), corresponding to 31 lead variants across 30 genomic loci. Estimated effective sample sizes were 234,013 for UKB CBP, 158,010 for CHARGE-UKB CBP, 437,112 for FinnGen dorsalgia, 409,477 for MVP dorsalgia, and 310,280 for UKB CMSKP. A summary of the number of significant single nucleotide polymorphisms (SNPs; P < 5×10⁻⁸), lead SNPs, genomic loci, heritability (h²), and genomic inflation factor (lambda GC) for each trait is detailed in **Supplementary Table 2**.

### SNP-based heritability and Genetic correlations with related traits

We used LD Score Regression (LDSC) to estimate the SNP-based heritability (*h*^2^) and pairwise genetic correlations (rg) between CBP in the UKB and four genetically related traits: CHARGE-UKB CBP, dorsalgia from the FinnGen study, dorsalgia from the Million Veteran Program, and UKB CMSKP. The SNP-based heritability estimates on the observed scale varied across traits, with the highest being the UKB CBP phenotype (*h*² = 0.1193, SE = 0.0053). Trait-specific heritability estimates were as follows: CHARGE-UKB CBP: h² = 0.0356 (SE = 0.0034); FinnGen dorsalgia: *h*² = 0.0535 (SE = 0.0022); MVP dorsalgia: *h*² = 0.0698 (SE = 0.0031); and CMSKP (UKB): *h*² = 0.0775 (SE = 0.0034) (Table 1). All traits exhibited acceptable LDSC intercepts (ranging from 1.0003 to 1.2587), indicating that inflation in the test statistics was primarily due to polygenicity rather than confounding bias. Each of the traits showed a strong and statistically significant genetic correlation with UKB CBP, with all rg values ∼ 0.7 (**Figure 1**).

**Figure 1.**
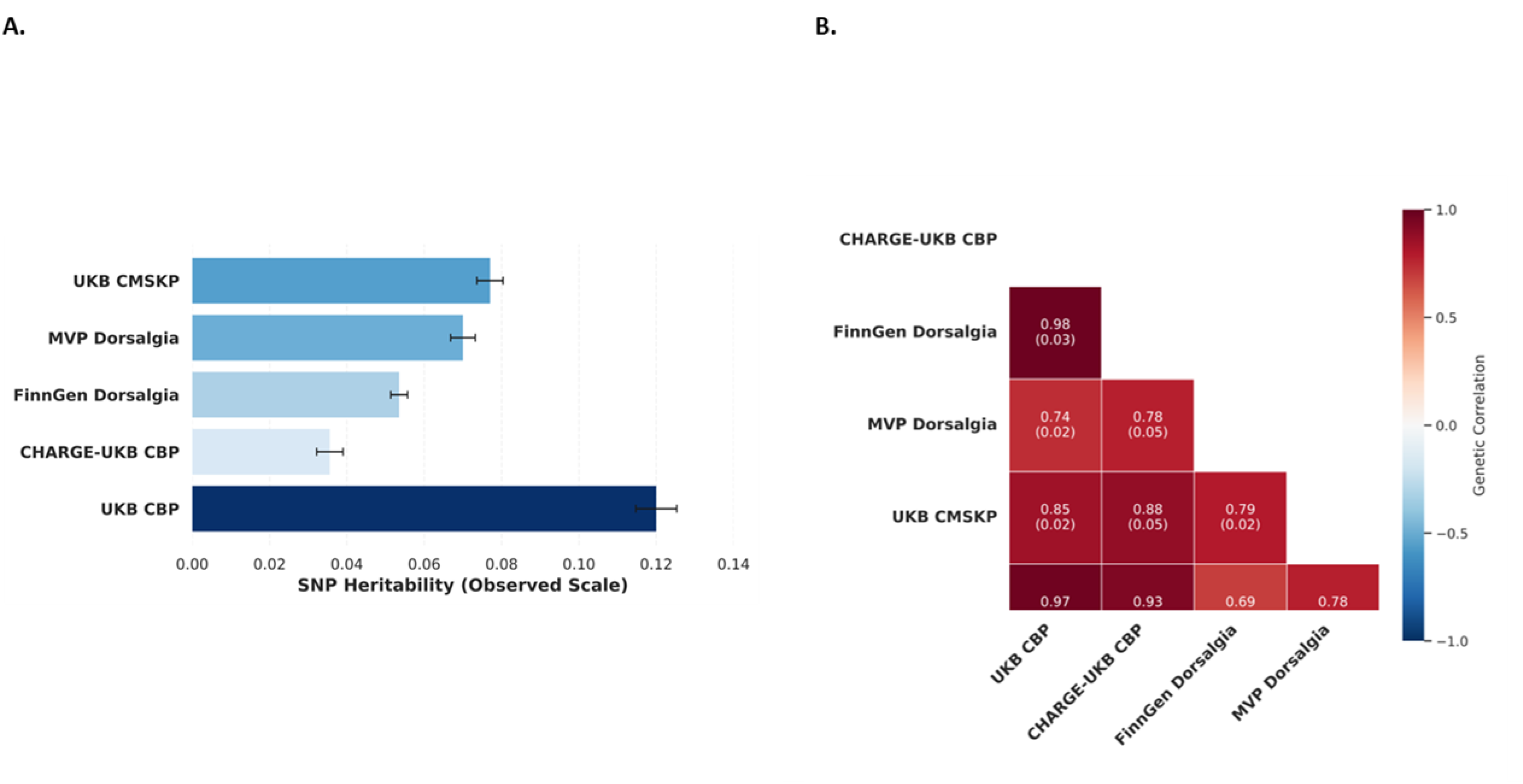
SNP-based heritability and genetic correlations across back pain-related traits. (**A.)** SNP-based heritability estimates (on the observed scale) for chronic back pain (CBP) in the UK Biobank, CBP from the CHARGE-UKB meta-analysis, dorsalgia phenotypes from FinnGen and MVP, and chronic musculoskeletal pain (CMSKP) from the UKB. Bars represent point estimates with standard errors. (**B**.) Genetic correlations (rg) between UKB CBP and related traits estimated using bivariate LD Score Regression. Values shown within cells indicate the genetic correlation and standard error. All correlations were statistically significant (p < 0.05).

**Table 1.** Predictive Performance of Polygenic Risk Scores (PRS) across Discovery (Base) and Validation (Target) datasets.

| Base | Target | Adjustment model | AUC | AUPRC | Nagelkerke R<sup>2</sup> | Brier Score |
| --- | --- | --- | --- | --- | --- | --- |
| UKB | CLSA | PRS only | 0.582 | 0.312 | 0.023 | 0.186 |
| UKB | CLSA | PRS + Age + Age<sup>2</sup> + Sex | 0.600 | 0.328 | 0.034 | 0.184 |
| UKB | CLSA | PRS + Age + Age<sup>2</sup> + Sex + PCs 1-10 | 0.604 | 0.330 | 0.038 | 0.184 |
| MTAG | CLSA | PRS only | 0.603 | 0.329 | 0.037 | 0.184 |
| MTAG | CLSA | PRS + Age + Age<sup>2</sup> + Sex | 0.617 | 0.343 | 0.047 | 0.183 |
| MTAG | CLSA | PRS + Age + Age<sup>2</sup> + Sex + PCs 1-10 | 0.621 | 0.346 | 0.051 | 0.182 |
| MTAG | CARTaGENE | PRS only | 0.615 | 0.29 | 0.041 | 0.164 |
| MTAG | CARTaGENE | PRS + Age + Age<sup>2</sup> + Sex | 0.636 | 0.329 | 0.061 | 0.161 |
| MTAG | CARTaGENE | PRS + Age + Age<sup>2</sup> + Sex + PCs 1-10 | 0.638 | 0.335 | 0.064 | 0.161 |
| MTAG | GENE-PAR+ CARTaGENE | PRS only | 0.670 | 0.415 | 0.102 | 0.171 |
| MTAG | GENE-PAR+ CARTaGENE | PRS + Age + Age<sup>2</sup> + Sex | 0.760 | 0.584 | 0.260 | 0.147 |
| MTAG | GENE-PAR+ CARTaGENE | PRS + Age + Age<sup>2</sup> + Sex + PCs 1-10 | 0.785 | 0.616 | 0.306 | 0.141 |

### Multi-trait analysis

Applying MTAG to these five summary statistics increased the mean chi-square (χ²) for the UKB CBP trait from 1.393 to 1.826, reflecting a genome-wide strengthening of association signals for CBP and indicating improved power and more precise estimation of SNP effects due to the integration of genetically correlated traits. The effective sample size for UKB CBP nearly doubled from 234,013 to 492,694 and the MTAG-boosted CBP phenotype showed an observed-scale h² of 0.2599 (SE = 0.0080). The MTAG weight factors revealed the strongest contribution from UKB CBP itself (1.416), followed by CMSKP (1.169), MVP dorsalgia (1.094), and FinnGen dorsalgia (1.062), with CHARGE-UKB CBP contributing modestly (0.786).

Across all five traits, a total of 5,482,652 overlapping SNPs were retained and used in the joint analysis. The MTAG-boosted analysis detected 390 independent significant SNPs in the UKB CBP, including 179 lead SNPs across 156 genomic loci (**Supplementary Table 2**).

To assess type I error control under the MTAG model, we examined the maximum false discovery rate (MaxFDR) for each trait. All traits exhibited low MaxFDR values, consistent with well-controlled false positive rates. Specifically, MaxFDR was 0.00107 for MTAG-boosted UKB CBP, 0.00148 for CHARGE-UKB CBP, 0.00286 for FinnGen dorsalgia, 0.00272 for MVP dorsalgia, and 0.00062 for CMSKP.

Among the 4,438 SNPs reaching genome-wide significance in the MTAG-boosted UKB CBP (excluding MVP), 4,429 (99.8%) were available in MVP dorsalgia. Of these, 3,544 (79.9%) exhibited concordant direction of effect and nominal replication significance (P < 0.05). Similarly, for the MTAG-boosted CHARGE-UKB CBP trait, 4,386 of 4,395 significant SNPs (99.8%) were present in MVP, with 3,502 (79.7%) meeting the same replication criteria.

### Polygenic risk score development

CBP PRS were derived in the CLSA using five-fold cross-validation to optimize LD clumping parameters and p-value thresholds. Across 45 PRS models (r² = 0.1, 0.5, 0.7; window = 250, 500, 1,000 kb), the best discrimination was achieved at r² = 0.5 and window = 1,000 kb (AUC = 0.6297 ± 0.0135, **Supplementary Table 3**). The corresponding PRS-only model yielded an AUC of 0.6028 ± 0.0115, and the optimal score included 82,768 ± 19 SNPs (range: 82,741-82,794). Each 1 SD increase in PRS was associated with an OR of 1.455 (95% CI: 1.394-1.518) for CBP. **Table 1** summarizes discrimination, explained variances, and overall model calibration for all PRS models.

In the CLSA, cases (n = 3,201) and controls (n = 9,484) had nearly identical mean ages (62.1 ± 9.8 vs. 62.2 ± 9.9 years; p = 0.488). The distribution of standardized PRS values in the CLSA revealed a rightward shift among cases compared to controls, indicating that individuals with CBP had higher polygenic scores on average. Cases had a mean PRS 0.40 SD higher than controls (p = 7.6×10⁻¹⁹) (**Figure 2A**). In the unadjusted model, the MTAG-PRS achieved an area under the ROC curve (AUC) of 0.603 (95% CI: 0.588–0.618; AUPRC = 0.329; Nagelkerke R² = 0.037) and a Brier score of 0.184, modestly higher than the UKB CBP-only PRS (AUC = 0.582; AUPRC = 0.312; R² = 0.023; Brier score = 0.186). Fully adjusting the model by adding age, age², sex, and PCs 1-10 increased the MTAG-PRS AUC to 0.617 (AUPRC = 0.343; R² = 0.047), versus 0.600 (AUPRC = 0.328; R² = 0.034) for the UKB-PRS. **Figure 2B** shows the ROC of the fully adjusted MTAG-PRS (AUC = 0.621; AUPRC = 0.346, **Figure 2C**; R² = 0.051), which was slightly better than that of UKB CBP-only PRS (AUC = 0.604; AUPRC = 0.330; R² = 0.038). The MTAG-boosted PRS explained an additional 2.15% of the variance in CBP beyond the covariate-only model, while the additional variance explained by the UKB CBP-only PRS was 1.34%. This corresponds to a 0.81% gain in predictive power. Individuals in the top 5% of the MTAG PRS distribution had more than twice the odds of reporting CBP compared to the middle 40-60% group (OR = 2.11; 95% CI: 1.75-2.53) (**Figure 2D**).

**Figure 2.**
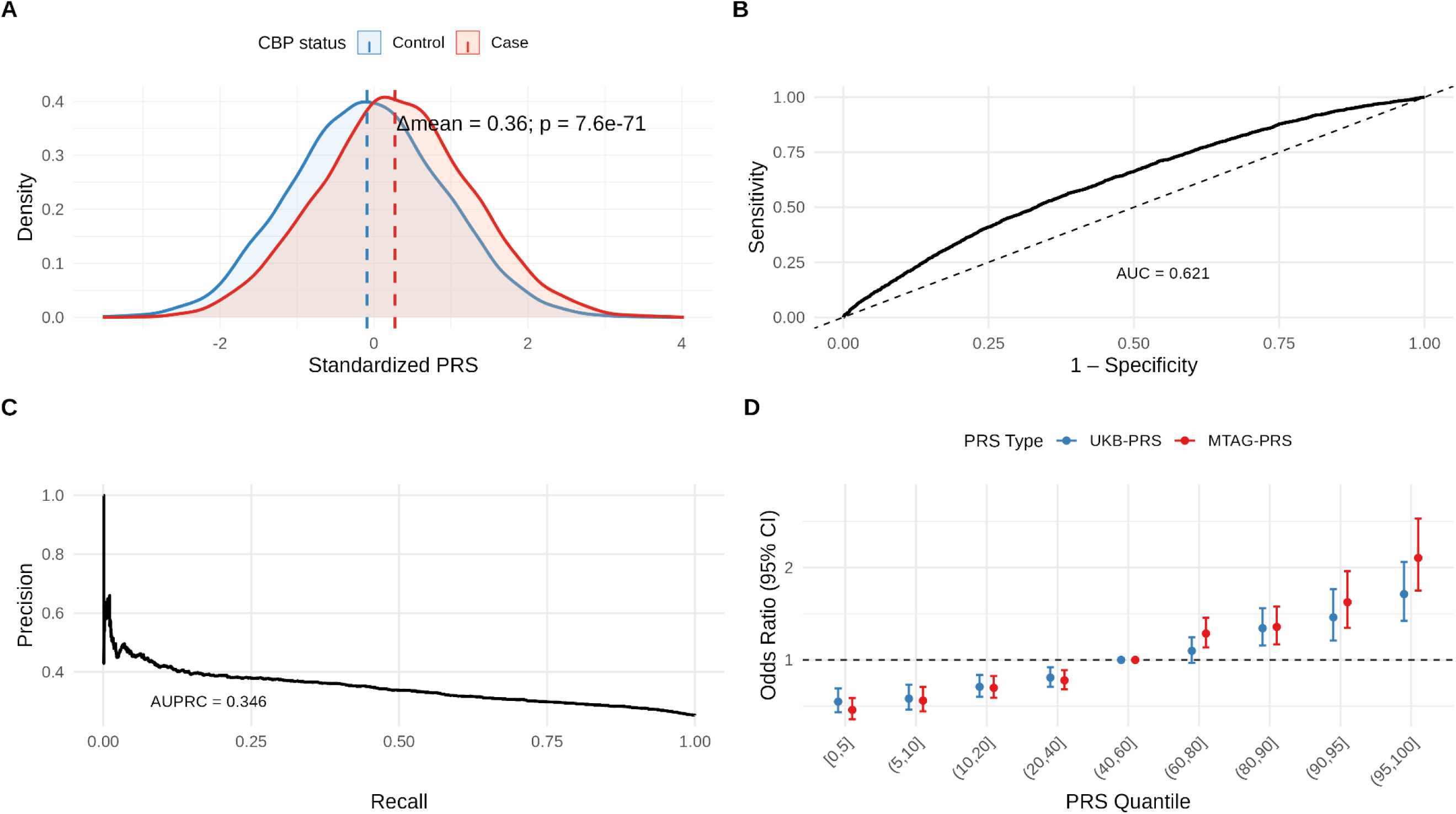
Internal Validation of Polygenic Risk Scores for Chronic Back Pain in the CLSA Cohort **(A.**) Density distribution of standardized PRS at optimized parameters (clump r2 =0.5; clump kb =1000; p < 0.1; 82,762 SNPs included) for all cases and controls. (Δmean = 0.36; P = 7.6× 10⁻^71^, Welch’s t-test). (**B**.) ROC curve for the fully adjusted model (AUC = 0.621). (**C**.) Precision-recall curve for the same model (AUPRC = 0.35). (**D**.) Odds ratios for chronic back pain by PRS quantile bin, comparing MTAG-PRS (red) and UKB-only PRS (blue). The middle quantile (40-60%) was used as the reference group (OR = 1). Error bars represent 95% confidence intervals.

### External PRS validation

Within the CARTaGENE – case-control sample (n = 2,951), CBP cases (n = 632) were significantly older than controls (n = 2,319), with mean ages of 56.98 ± 7.85 versus 54.89 ± 8.05 years, respectively (p = 4.62 × 10⁻⁹). The standardized PRS was significantly higher in cases than controls with a mean difference of 0.40; P = 1.7× 10⁻^19^ (**Supplementary Figure 3A**). The MTAG-PRS achieved an AUC = 0.615 (AUPRC = 0.290; R² = 0.041; Brier score = 0.164) in the unadjusted model, increasing to AUC = 0.636 (AUPRC = 0.329; R² = 0.061; Brier score = 0.161) with demographics and to AUC = 0.638 (AUPRC = 0.335; R² = 0.064; Brier score = 0.161) after full adjustment (**Table 1, Supplementary Figure 3B, Supplementary Figure 3C**).

In the combined GENE-PAR + CARTaGENE – controls dataset, the overall mean age was 56.3 ± 9.1 years (n = 1,152), and this remained identical (56.3 ± 9.1 years) after excluding eight participants with missing covariate data (n = 1,144). Cases were significantly older than controls (60.6 ± 10.3 vs. 54.8 ± 8.2 years; p = 2.8 × 10⁻¹⁶). The standardized PRS was significantly higher in cases than controls (0.441 ± 2.58 vs −0.147 ± 2.40; p = 7.91 × 10⁻¹⁷; **Figure 3A**). The MTAG-PRS alone yielded AUC = 0.670 (AUPRC = 0.415; R² = 0.102) (**Table 1**). Demographics adjustment raised the AUC to 0.760 (AUPRC = 0.584; R² = 0.260), and the fully adjusted model achieved AUC = 0.785 (AUPRC = 0.616; R² = 0.306) (**Figure 3B; Figure 3C**).

**Figure 3.**
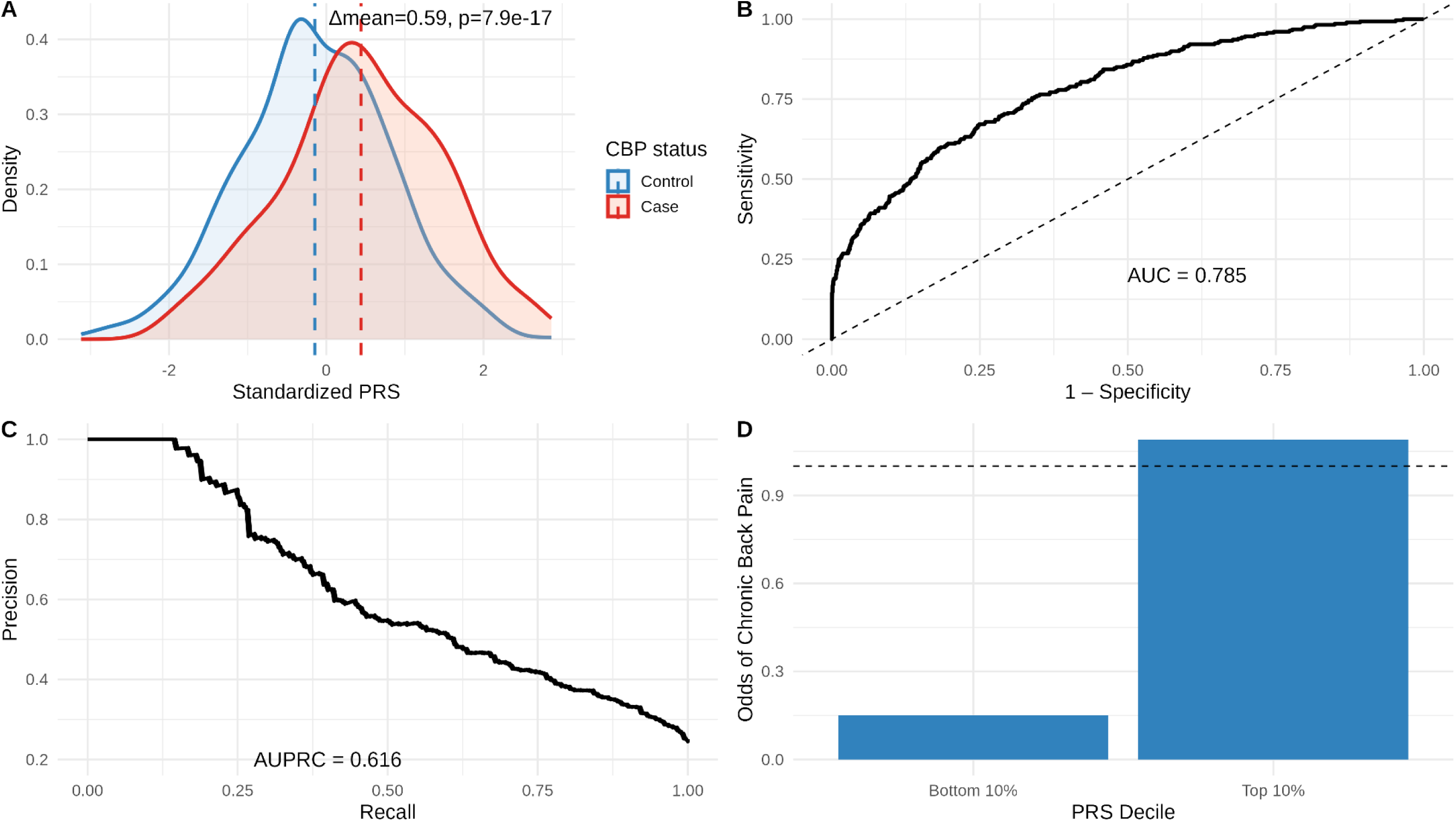
External validation of the chronic back pain polygenic risk score (PRS) in GENE-PAR+CARTAGENE cohort (**A.**) Density of standardized PRS in controls (blue) and cases (red), with dashed lines at group means (Δmean = 0.59; P = 7.9× 10⁻^17^, Welch’s t-test). (**B.)** ROC curve for the fully adjusted model (AUC = 0.785). (**C.)** Precision–recall curve for the same model (AUPRC = 0.62). (**D**.) Odds of chronic back pain GENE-PAR participants present in the top vs bottom 10% of the PRS distribution (OR = 6.70, 95% CI: 3.60-13.10).

### Clinical PRS Validation

When we stratified the GENE-PAR + CARTaGENE – controls by PRS decile (n = 116 per decile), 51.7% of individuals in the top decile (60 of 116) were cases, compared with 13.8% (16 of 116) in the bottom decile. This enrichment corresponded to an OR of 6.64 (95% CI 3.60-13.10; p = 7.88 × 10⁻¹⁰) for top versus bottom decile (**Figure 3D**). Clinical and psychosocial characteristics of the 76 participants in the GENE-PAR dataset in the extreme PRS deciles (top decile, n = 60; bottom decile, n = 16) are presented in **Supplementary Table 4.** Participants in the top decile had significantly greater pain interference (65.46 ± 5.73 vs 60.94 ± 6.94; p = 0.026), greater pain intensity (6.33 ± 1.95 vs 4.69 ± 2.39; p = 0.022), were more likely to report pain catastrophizing (66.7% vs 31.2%; p = 0.02), and reported a greater burden of somatic symptoms (SCL “no-pain” sub-scale = 1.57 ± 0.82 vs 1.12 ± 0.72; p = 0.040) compared with those in the bottom decile (**Table 2**). Seemingly counterintuitively, participants in the top PRS decile also showed better physical function (mean ± SD 38.3 ± 5.8 vs 35.3 ± 4.8; p = 0.047).

**Table 2.** Subset of Clinical and Psychosocial Characteristics of GENE-PAR participants in the top and bottom 10% of the PRS distribution.

|  | **Bottom 10% PRS**<br>**(n = 16)** | **Top 10% PRS**<br>**(n = 60)** | **P-value\*** |
| --- | --- | --- | --- |
| **Pain intensity, mean**<br>**(SD)** | 4.69 (2.39) | 6.33 (1.95) | **0.019** |
| Missing | 0 | 0 |  |
| **Pain interference,**<br>**mean (SD)** | 60.94 (6.94) | 65.46 (5.73) | **0.026** |
| Missing | 0 | 0 |  |
| **Catastrophizing, n**<br>**(%)** |  |  |  |
| No (ref) | 11 (68.80) | 20 (33.30) | **0.020** |
| Yes | 5 (31.20) | 40 (66.70) |  |
| Missing | 0 | 0 |  |
| **Physical function,**<br>**mean (SD)** | 35.27 (4.83) | 38.34 (5.81) | **0.047** |
| Missing | 1 | 4 |  |
| **Somatization**<br>**(without pain), mean**<br>**(SD)** | 1.12 (0.72) | 1.57 (0.82) | **0.040** |
| Missing | 0 | 0 |  |

## Discussion

We demonstrated that integrating genetically correlated traits via MTAG enhances PRS performance for CBP. By combining five European-ancestry GWAS (UKB CBP, CHARGE-UKB CBP, FinnGen dorsalgia, MVP dorsalgia, and UKB CMSKP) and accounting for overlapping samples, we nearly doubled the sample size for CBP cases (going from 234,013 to 492,717) and more than doubled its SNP-heritability (going from 11.9% to 26.0% on the observed scale). MTAG identified 156 independent CBP loci, more than fivefold the 30 detected with the UKB CBP-only data. Incorporating genetically correlated phenotypes, namely back pain without a duration criterion (dorsalgia) and chronic musculoskeletal pain, defined using the same duration threshold as the CBP primary trait at 3 or more months across multiple musculoskeletal sites, increased power for otherwise subthreshold CBP signals.

Translating these MTAG-derived discoveries into PRS development, we observed substantial improvements in predictive performance across both population-based and clinician-ascertained samples, despite differences in phenotype definitions and sample composition. The larger effective sample size and increased power from MTAG enabled the inclusion of more informative SNPs with stronger effect estimates, which collectively enhanced the ability of PRS to discriminate CBP cases from controls and explained a greater proportion of phenotypic variance. In contrast, earlier CBP GWAS identified only three loci *(SOX5, CCDC26/GSDMC, DCC*) in ∼158,000 Europeans[69], and subsequent meta-analyses conducted by CBP consortia expanded this number to just over 100 loci [67]. Given CBP’s extreme polygenicity, discovery improves via larger samples and by leveraging cross-trait genetic correlation via MTAG to achieve locus-specific boosts in power where trait effects align.

Despite substantial discovery of CBP loci, the predictive performance of CBP PRS remains modest; to date, the only prediction-focused study reported an AUC of 0.56 evaluated in large-scale population-based samples[73]. In the CLSA, our MTAG-derived PRS produced a slight improvement in AUC (0.603 vs. 0.582), AUPRC (0.329 vs. 0.312), and Nagelkerke’s pseudo-R² (0.037 vs. 0.023) compared to the UKB-only PRS. After adjusting for age, sex, and the first 10 principal component vectors, discrimination rose to AUC = 0.621 (R² = 0.051) versus 0.604 (R² = 0.038) for the UKB CBP-only score. Although numerically small, an absolute AUC gain of ∼0.02 represents a tangible advance in risk stratification for CBP, yielding more high-risk individuals correctly identified at matched specificity. In fact, the MTAG-derived PRS exceeded the ∼0.003 AUC increase produced by an improved knee osteoarthritis PRS[54]. Brier score improved only marginally (0.184 vs. 0.186; fully adjusted = 0.182). Importantly, in the CARTaGENE-only analysis, the fully adjusted model also showed gains (AUC = 0.638, AUPRC = 0.335, R² = 0.064, Brier = 0.161), although largely less substantial and similar to those seen in the CLSA.

External validation in the dataset including clinician-ascertained CLBP cases (GENE-PAR) and CARTaGENE controls yielded substantially greater performance. According to the framework proposed by Swets (1988) for evaluating the clinical utility of predictive tests[70], our MTAG-PRS alone approached the levels of clinical utility (AUC > 0.7), displaying an AUC of 0.670 (R² = 0.102). Following full adjustment, the AUC surpassed the level of potential clinical utility (0.785; R² = 0.306), an unprecedented finding in chronic pain. Brier score also showed an improvement compared to that obtained in the CLSA (Brier = 0.141 vs. 0.182 in CLSA), indicating better overall accuracy of the predicted probabilities. Combined, these findings demonstrate that CBP PRS benefits substantially from clinical characterization of CBP status.

A crucial determinant of PRS accuracy is discovery GWAS sample size: larger training datasets reduce standard errors and sharpen SNP-effect estimates[3; 57]. Leveraging genetically correlated traits (e.g., via MTAG or multi-PGS) further enlarges effective sample size and improves prediction[3]. These gains also appear when training on broader phenotypes, where increased *N* boosts PRS performance despite greater phenotypic heterogeneity. The corollary is a trade-off between shallow phenotyping (large N, lower specificity) and deep phenotyping (smaller N, higher specificity). The trade-off between sample size and extent of phenotyping was recently explored in the context of major depressive disorder (MDD) for the base dataset. Using large, minimally phenotyped meta-analysis GWAS summary data (∼246k cases) yielded stronger PRS associations with clinically diagnosed major depressive disorder (MDD) (OR≈1.75 per SD) than a much smaller strictly diagnosed GWAS (OR≈1.14); however, when sample size was equalized, the clinically defined GWAS produced the most specific predictor for clinical MDD[53]. When considering the target dataset, other studies of various health outcomes indicated that PRSs perform better in rigorously phenotyped samples[51; 77]. Consistent with prior findings in other phenotypes, while enlarging the base GWAS improved locus discovery and slightly increased PRS performance, our findings showed that CBP-specific clinical phenotyping in the target dataset drove a substantial increase in the CBP-PRS performance. Another non-exclusive explanation for higher PRS performance in the target dataset is that recruitment for the GENE-PAR Study may have captured individuals with more severe chronic back pain, as they may have been more prone to respond to an invitation to participate. This could enrich genetic liability and inflate apparent PRS performance, constituting ascertainment bias due to severity enrichment. Despite this, GENE-PAR likely aligns well with the intended target population for a clinical setting PRS application, which is the downstream goal of this work.

In addition, in this combined dataset, (GENE-PAR + CARTaGENE – controls), participants spanned a wide age range, and CBP cases were on average substantially older than controls, creating a clear age-related imbalance. Given that CBP prevalence typically follows an “inverted-U” trajectory - rising through mid-life, peaking in the early 60s, and declining thereafter[35; 36], incorporating both linear and quadratic age terms captured this non-linear risk pattern and meaningfully improved model discrimination. By contrast, in CLSA, where participants were older and the age distribution was narrower, there was no significant difference in age between cases and controls, and including age² contributed little additional predictive value. The CARTaGENE subset, which had intermediate age variability, showed smaller but still detectable gains when modeling age non-linearly. Together, these findings suggest that accounting for non-linear age effects enhances prediction in samples with broader age distributions, whereas its impact is minimal in older, more homogeneous populations.

PRS decile analysis in the GENE-PAR + CARTaGENE controls also showed that individuals in the top decile had sixfold higher CBP odds versus those in the bottom decile. Notably, among GENE-PAR CLBP cases, top-decile individuals reported greater pain intensity, pain interference, catastrophizing, and a greater burden of somatic symptoms. These measures have consistently been shown to inform cLBP outcomes, including the risk of chronicity[34; 56]. Pragmatically, our findings suggest that PRSs can be used to stratify patients and ultimately inform targeted allocation of healthcare resources or early intervention strategies to minimize the burden of chronic pain. Despite the abovementioned greater burden of CLBP symptoms, participants in the highest PRS decile displayed better physical function. This has been reported before and suggests that these individuals develop adaptive mechanisms of coping or resilience that enable them to remain active despite a heightened symptom burden[50; 56].

Our study has limitations that must be considered. MTAG assumes a single, genome-wide covariance structure of SNP effects; loci with trait-specific architectures may be misestimated. PRS are ancestry-dependent and portability of PRS using a European base population on African ancestry target populations is particularly poor[49]. The method proposed here can be extended to other ancestries as GWAS of CBP become available. In this study, we constructed PRS via clumping-and-thresholding because it is computationally efficient and enables the systematic exploration of multiple tuning parameters across large-scale datasets. However, other approaches that model linkage disequilibrium more comprehensively have shown improved predictive performance in other complex traits[31; 55; 61; 78]. Future studies could evaluate whether such strategies further enhance CBP risk prediction within the MTAG framework. Finally, despite significant improvements, our PRS explains only a fraction of CBP heritability, reflecting the influence of environmental, psychosocial, and gene-environment interactions; thus, PRS should complement, not replace, established clinical and lifestyle risk factors in comprehensive CBP risk models.

Our study benefits from several methodological strengths. By integrating five genetically correlated pain-related traits using MTAG, we increased discovery power without requiring additional individual-level data. Independent GWAS replication, excluding MVP dorsalgia from the MTAG analysis, showed that ∼80% of lead SNPs remained directionally consistent and nominally significant in MVP, supporting the robustness of our findings. We further optimized PRS construction through five-fold cross-validation in CLSA, minimizing overfitting and improving parameter selection. Finally, external validation in both population-based and clinician-ascertained samples demonstrated the generalizability of our MTAG-derived PRS.

In summary, our study demonstrates that integrating genetically correlated traits through MTAG substantially enhances discovery and predictive performance for chronic back pain. By increasing effective sample size and SNP-heritability, we achieved unprecedented levels of PRS discrimination, particularly in clinically characterized samples. These findings establish a proof-of-principle that multi-trait approaches, coupled with rigorous phenotyping, can move CBP PRS toward thresholds of clinical utility. Continued expansion across diverse ancestries, adoption of advanced statistical methods, and integration with clinical and environmental factors will be essential to translate these genetic insights into tools that improve prevention, risk stratification, and personalized management of chronic back pain.

## Data Availability

The data used in this study were obtained from multiple cohorts. UK Biobank data are available to bona fide researchers through application to the UK Biobank
). CLSA data are accessible to qualified researchers through application to the Canadian Longitudinal Study on Aging
). CARTaGENE data are available through application to CARTaGENE. FinnGen summary statistics are publicly available through the FinnGen project (https://www.finngen.fi/en
). MVP summary statistics are available at https://mvp-ukbb.finngen.fi
. CHARGE-UKB meta-analysis summary statistics are available at the GWAS Archive (https://gwasarchive.org
) under "Chronic back pain" Summary statistics generated in this study are available from the corresponding author upon reasonable request.

## ACKNOWLEDGMENTS

The authors would like to acknowledge the Canadian Institutes of Health Research (CIHR PJT 190092), that have funded this study. The authors also would like to acknowledge the LAEF for the LAEF PhD Studentship awarded to R.O. The authors have no conflict of interest to declare.

The authors thank the participants and investigators of data sources used in this study, including the UK Biobank resource (application #20802), the CLSA (application #190213), the FinnGen study, the MVP, the CHARGE and PainOmics consortia, the CARTaGENE and GENE-PAR.

## Notes

### Competing Interest Statement

The authors have declared no competing interest.

### Funding Statement

This study was supported by the Louise and Alan Edwards Foundation (LAEF) through a Grant in Pain Research to C.B.M and a Studentship in Pain Research to R.O., and by a Canadian Institutes of Health Research (CIHR) Project Grant to C.B.M and A.V.G [CIHR PJT 190092]. The authors and their institutions did not receive payment or services from a third party for any aspect of the submitted work beyond these sources of funding

### Author Declarations

Ethics Committee of McGill University gave ethical approval for this work (UK Biobank approval #A03-M20-21B; CLSA approval #A05-M25-19A)

