## Supplementary Information for "A multi-trait approach improves polygenic risk scores for chronic back pain across population-based and clinically ascertained samples"

[1]Faculty of Dental Medicine and Oral Health Sciences, McGill University, QC, Canada.
[2]The Alan Edwards Centre for Research on Pain, McGill University, QC, Canada.
[3]Department of Anesthesia, Faculty of Medicine and Health Sciences, McGill University, Montreal, QC, Canada.
[4]Department of Human Genetics, McGill University, QC, Canada.
[5] Canada Excellence Research Chair in Genomic Medicine.
[6] Victor Phillip Dahdaleh Institute of Genomic Medicine at McGill University

**Supplementary Figures**

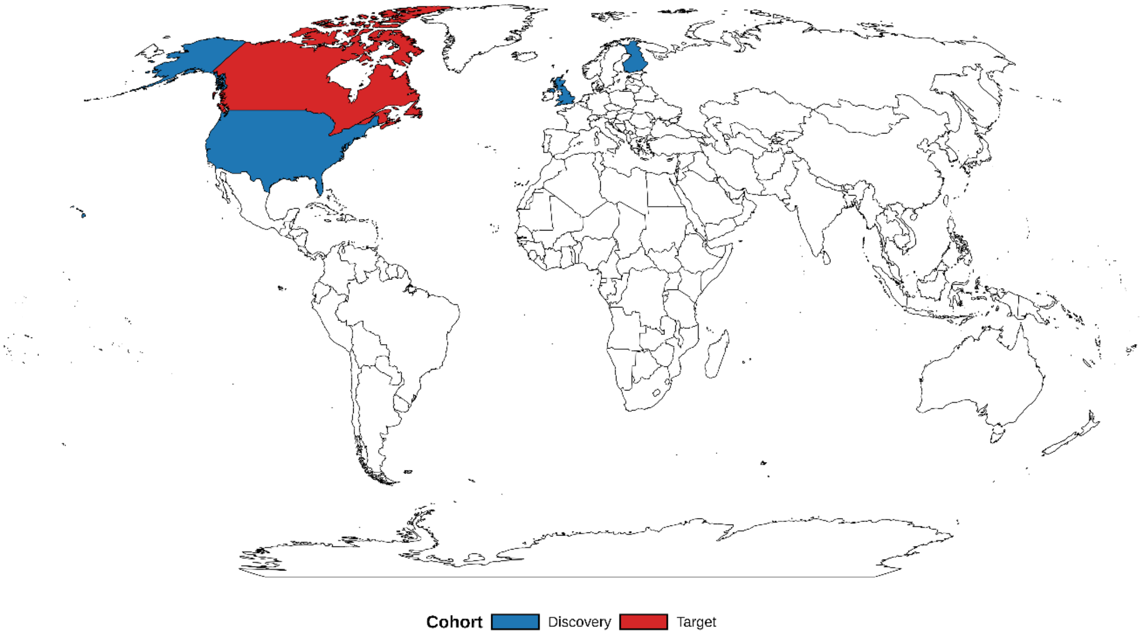


**Supplementary Figure 1**. Geographic distribution of cohorts included in the multi-trait GWAS and polygenic risk score (PRS) analyses of chronic back pain. Colors represent discovery cohorts (UK Biobank, CHARGE-UKB, FinnGen, and MVP) and target cohorts (CLSA and GENE-PAR+CARTaGENE). All cohorts consist of participants of European ancestry.

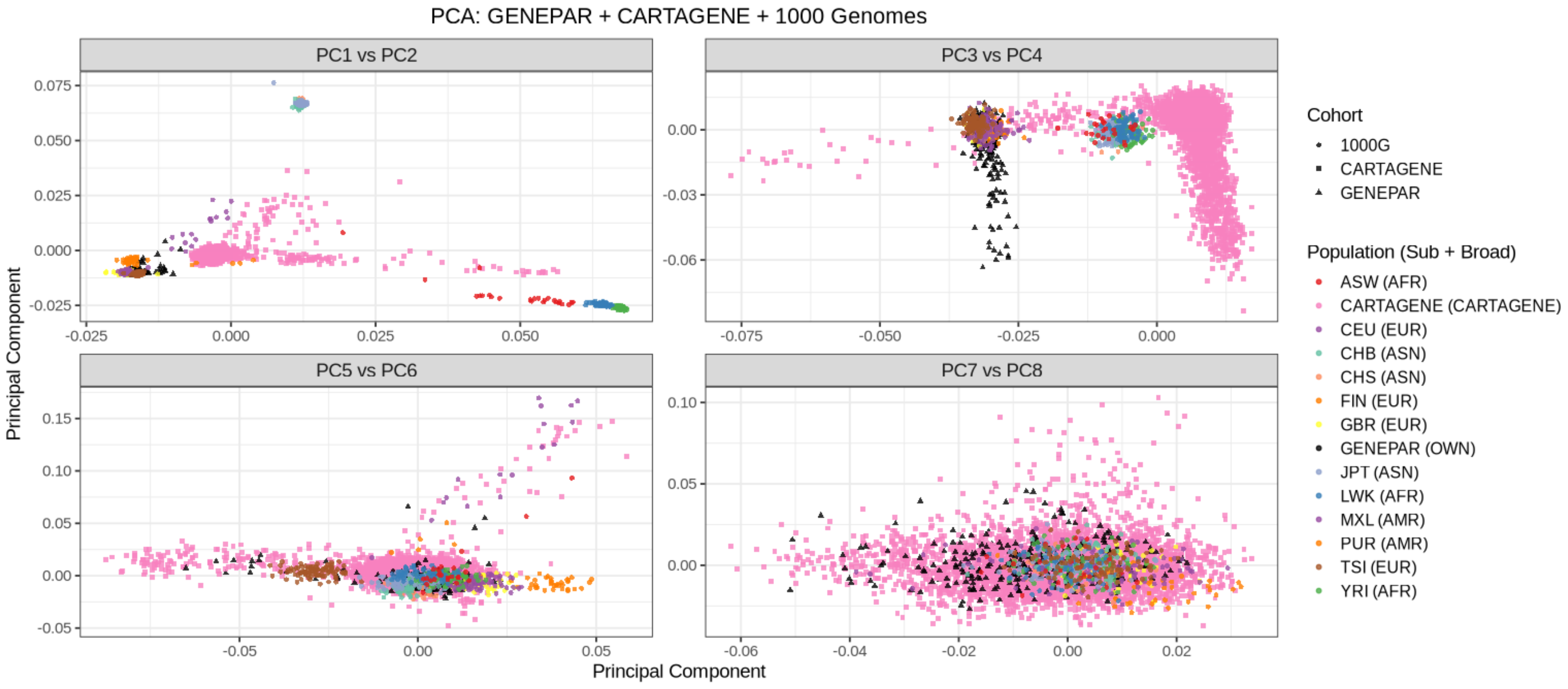


**Supplementary Figure 2**. Principal component analysis (PC1-PC8) situating GENEPAR and CARTaGENE participants within the ancestry clusters defined by 1000 Genomes reference population.

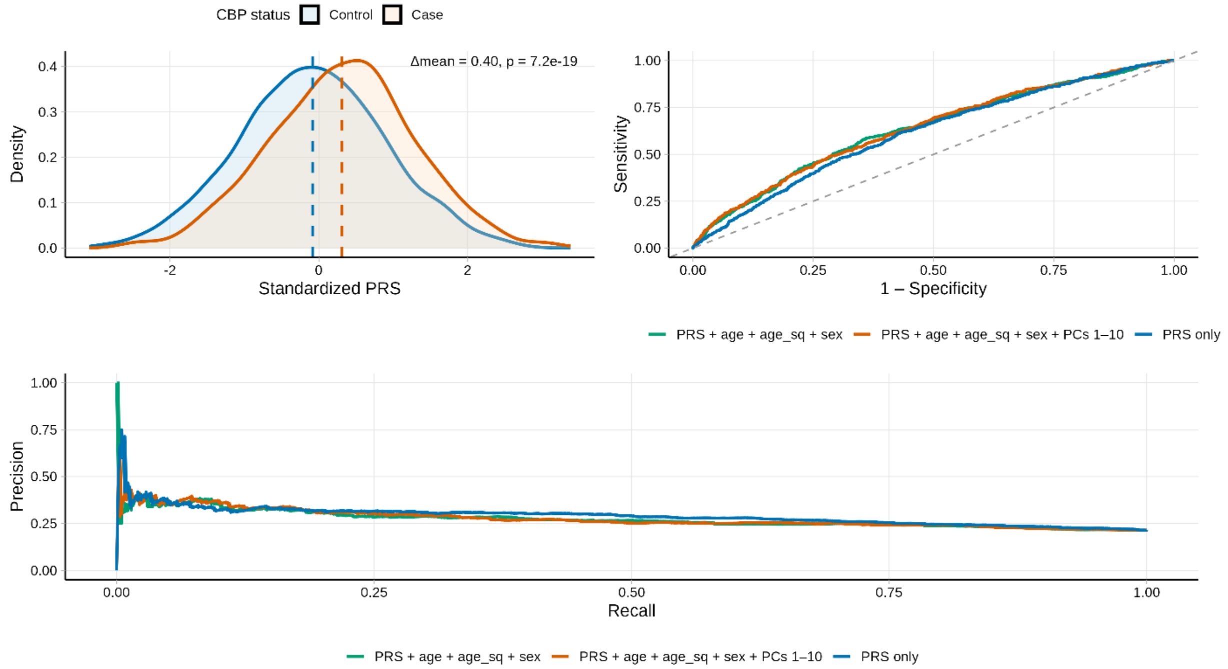


**Supplementary Figure 3**. External validation of the chronic back pain polygenic risk score (PRS) in CARTaGENE cohort (**A.**) Density of standardized PRS in controls (blue) and cases (red), with dashed lines at group means (Δmean = 0.40; P = $1.7\times 10^{-19}$, Welch's t-test). (**B.)** ROC curve for the fully adjusted model (AUC = 0.638. (**C.)** Precision-recall curve for the same model (AUPRC = 0.34).

**Supplementary Table 1.** Description of datasets used for discovery and target in the construction of a polygenic risk score (PRS) for chronic back pain. N stands for the number of participants of European ancestry.

| **Trait** | **Role** | **Total (N)** | **Cases (N)** | **Cohort** | **Phenotype definition** |
|---|---|---|---|---|---|
| Chronic back pain (CBP) | Discovery (base) | 234,013 | 70,643 | United Kingdom Biobank | Self-reported, back pain more than 3 months |
| Chronic back pain (CBP) | Discovery (base) | 158,010 | 29,531 | United Kingdom Biobank, Cohorts for Heart and Aging Research in Genomic Epidemiology (CHARGE) consortium (PMC6159857) | Self-reported, back pain more than 3 months |
| Dorsalgia | Discovery (base) | 437,112 | 83,888 | FinnGen Release 12 | Electronic Health Record (EHR), International Statistical Classification of Diseases, 10th Revision (ICD-10) code M54 |
| Dorsalgia | Discovery (base) | 409,477 | 221,759 | Million Veteran Program (MVP) | Electronic Health Record (EHR), International Statistical Classification of Diseases, 10th Revision (ICD-10) code M54 |
| Chronic musculoskeletal pain (CMSKP) | Discovery (base) | 310,280 | 156,235 | United Kingdom Biobank | Self-reported, neck/shoulder + knee pain + hip + back pain more than 3 months |
| Chronic back Pain | Target | 12685 | 3,201 | Canadian Longitudinal Study on Aging (CLSA) | Self-reported back pain for more than 3 months |
| Chronic back Pain | Target | 3183 | 632 | CARTaGENE | Self-reported history of physician-diagnosed CBP |
| Chronic back Pain | Target | 288 | 288 | GENE-PAR | Specialist-diagnosed LBP ≥3 months and present ≥½ of days in past 6 months |

**Supplementary Table 2**. Statistical summary of GWAS and MTAG results. Metrics include number of significant SNPs ($P < 5\times10^{-8}$), lead SNPs, genomic loci, heritability ($h^2$), lambda GC, and sample sizes for each cohort.

| | **UKB CBP** | | **CHARGE_UKB CBP** | | **FINNGEN DORS** | | **MVP DORS** | | **UKB CMSKP** | |
|---|---|---|---|---|---|---|---|---|---|---|
| | GWAS | MTAG | GWAS | MTAG | GWAS | MTAG | GWAS | MTAG | GWAS | MTAG |
| SigSNPs | 52 | 390 | 1 | 399 | 118 | 315 | 144 | 383 | 54 | 320 |
| Lead SNPs | 31 | 179 | 1 | 185 | 46 | 136 | 75 | 164 | 28 | 153 |
| Genomic locus | 30 | 156 | 1 | 161 | 31 | 120 | 71 | 149 | 26 | 138 |
| $\chi^2$ | 1.393 | 1.826 | 1.094 | 1.822 | 1.368 | 1.712 | 1.379 | 1.816 | 1.388 | 1.733 |
| $h^2$ | 0.119 | 0.2599 | 0.036 | 0.328 | 0.054 | 0.1078 | 0.069 | 0.125 | 0.078 | 0.155 |
| Lambda GC | 1.403 | 1.691 | 1.105 | 1.687 | 1.507 | 1.585 | 1.765 | 1.695 | 1.425 | 1.615 |
| Intercept | 1.023 | 0.952 | 1.0003 | 0.949 | 1.144 | 0.905 | 1.259 | 0.945 | 1.043 | 0.916 |
| Ratio | 0.047 | < 0 | 0.003 | < 0 | 0.240 | < 0 | 0.311 | < 0 | 0.084 | < 0 |
| N Mapped genes | 65 | 429 | 1 | 473 | 218 | 393 | 168 | 474 | 95 | 366 |
| Sample size | 234013 | 492717 | 158010 | 1388397 | 437112 | 846633 | 409477 | 880,839 | 310280 | 585594 |
| Overlapped SNPS | 5482652 | | 5482652 | | 5482652 | | 5482652 | | 5482652 | |

**Supplementary Table 3.** Five-fold cross-validated performance of CBP polygenic risk scores across LD clumping parameters in CLSA, including SNP counts.

| Clump r2 | Window size (kb) | Folds (N) | AUC (Full) mean ± SD | AUC (PRS-only) mean ± SD | SNPs in PRS (mean ± SD) | SNPs in PRS (range) | Pooled OR per 1 SD PRS (95% CI) |
|---|---|---|---|---|---|---|---|
| 0.1 | 250 | 5 | 0.6237 ± 0.0158 | 0.5964 ± 0.0123 | 56,686 ± 43,586 | 25,611–116,703 | 1.410 (1.352–1.471) |
| 0.1 | 500 | 5 | 0.6258 ± 0.0168 | 0.5995 ± 0.0149 | 26,946 ± 5,154 | 24,633–36,166 | 1.421 (1.362–1.482) |
| 0.1 | 1000 | 5 | 0.6254 ± 0.0163 | 0.5986 ± 0.0141 | 26,716 ± 5,120 | 24,417–35,875 | 1.416 (1.357–1.476) |
| 0.5 | 250 | 5 | 0.6294 ± 0.0138 | 0.6025 ± 0.0116 | 83,682 ± 17 | 83,657–83,703 | 1.453 (1.393–1.517) |
| 0.5 | 500 | 5 | 0.6297 ± 0.0137 | 0.6028 ± 0.0118 | 82,888 ± 18 | 82,863–82,914 | 1.455 (1.394–1.518) |
| **0.5*** | **1000** | **5** | **0.6297 ± 0.0135** | **0.6028 ± 0.0115** | **82,768 ± 19** | **82,741–82,794** | **1.455 (1.394–1.518)** |
| 0.7 | 250 | 5 | 0.6282 ± 0.0133 | 0.6001 ± 0.0106 | 222,384 ± 172,373 | 114,236–525,229 | 1.446 (1.385–1.510) |
| 0.7 | 500 | 5 | 0.6283 ± 0.0133 | 0.6002 ± 0.0109 | 208,569 ± 178,198 | 113,687–523,380 | 1.446 (1.386–1.510) |
| 0.7 | 1000 | 5 | 0.6283 ± 0.0131 | 0.6002 ± 0.0107 | 208,479 ± 178,145 | 113,624–523,195 | 1.446 (1.386–1.510) |

**Supplementary Table 4**. Clinical and psychosocial Characteristics of GENE-PAR participants in to and bottom 10% of PRS distribution.

| | **Bottom 10% PRS (n = 16)** | **Top 10% PRS (n = 60)** | ***P*-value*** |
|---|---|---|---|
| **Age (years),** mean (SD) | 62.60 (8.24) | 59.83 (9.85) | 0.275 |
| Missing | 1 | 0 | |
| **Sex,** n (%) | | | |
| Female | 8 (53.30) | 30 (50.00) | 1.000 |
| Male | 7 (46.70) | 30 (50.00) | |
| Missing | 1 | 0 | |
| **Race, n (%)** | | | |
| White | 15 (100.00) | 57 (98.28) | 1.00 |
| Non-white | 0 (0.00) | 1 (1.72) | |
| Missing | 1 | 2 | |
| **Education**, n (%) | | | |
| ≤ High school | 6 (37.50) | 25 (41.70) | 1.00 |
| > High school | 10 (62.50 | 35 (58.30) | |
| Missing | 0 | 0 | |
| **Employment**, n (%) | | | |
| Not employed | 11 (76.47) | 46 (80.70) | 0.736 |
| Employed | 4 (23.53) | 13 (19.30) | |
| Missing | 1 | 1 | |
| **BMI**, mean (SD) | 28.04 (7.13) | 30.90 (6.31) | 0.298 |
| Missing | 0 | 0 | |
| **History of cigarette use,** n (%) | | | |
| No (ref) | 14 (87.50) | 41 (68.33) | |
| Yes | 2 (12.50) | 19 (31.67) | 0.208 |
| Missing | 0 | 0 | |
| **Pain intensity,** mean (SD) | 4.69 (2.39) | 6.33 (1.95) | **0.019** |
| Missing | | | |
| **Sciatica pain,** n (%) | | | |
| No/not sure (ref) | 6 (37.50) | 11 (18.33) | 0.173 |
| Yes | 10 (62.50) | 49 (81.67) | |
| Missing | 0 | 0 | |
| **Pain comorbidities,** n (%) | | | |
| None (ref) | 8 (50.00) | 17 (28.3) | 0.180 |
| 1 or more | 8 (50.00) | 43 (71.7) | |
| Missing | 0 | 0 | |

| | | | |
|---|---|---|---|
| **Pain interference,** mean (SD) | 60.94 (6.94) | 65.46 (5.73) | **0.026** |
| Missing | 0 | 0 | |
| **Ongoing LBP treatments** | | | |
| **Opioids,** n (%) | | | |
| No (ref) | 4 (25.00) | 7 (11.70) | 0.229 |
| Yes | 12 (75.00) | 53 (88.30) | |
| Missing | 0 | 0 | |
| **Infiltration/injection,** n (%) | | | |
| No (ref) | 4 (25.00) | 13 (22.03) | 0.749 |
| Yes | 12 (75.00) | 46 (77.97) | |
| Missing | 0 | 1 | |
| **Exercise therapy**, n (%) | | | |
| No (ref) | 4 (26.67) | 15 (25.42) | 1.00 |
| Yes | 11 (73.33) | 44 (74.58) | |
| Missing | 1 | 1 | |
| **Psychological counselling**, n (%) | | | |
| No (ref) | 13 (81.25) | 40 (68.97) | 0.532 |
| Yes | 3 (18.75) | 18 (31.03) | |
| Missing | 0 | 2 | |
| **LBP-related surgery,** n (%) | | | |
| No (ref) | 10 (62.50) | 37 (61.70) | 1.00 |
| Yes | 6 (37.50) | 23 (38.30) | |
| Missing | 0 | 0 | |
| **Work absenteeism,** n (%) | | | |
| No (ref) | 2 (66.67) | 7 (38.89) | 0.553 |
| Yes | 1 (33.33) | 11 (61.11) | |
| Does not apply | 11 | 36 | |
| Missing | 2 | 6 | |
| **Physical function,** mean (SD) | 35.27 (4.83) | 38.34 (5.81) | **0.047** |
| Missing | 1 | 4 | |
| **Depression,** mean (SD) | 57.34 (6.27) | 59.59 (10.45) | 0.282 |
| Missing | 0 | 0 | |
| **Sleep disturbance,** mean (SD) | 51.43 (4.51) | 49.88 (4.84) | 0.240 |
| Missing | 0 | 0 | |
| **Catastrophizing,** n (%) | | | |
| No (ref) | 11 (68.80) | 20 (33.30) | **0.020** |
| Yes | 5 (31.20) | 40 (66.70) | |

| | | | |
|---|---|---|---|
| Missing | 0 | 0 | |
| **Kinesiophobia,** n (%) | | | |
| No (ref) | 8 (50.00) | 43 (71.70) | 0.136 |
| Yes | 8 (50.00) | 17 (28.30) | |
| Missing | 0 | 0 | |
| **Alcohol/drug overuse,** n (%) | | | |
| Never/rarely (ref) | 13 (86.67) | 50 (83.33) | 1.00 |
| Sometimes/often | 2 (13.33) | 10 (16.67) | |
| Missing | 1 | 0 | |
| **Neuropathic pain,** mean (SD) | 3.00 (1.67) | 3.88 (1.78) | 0.076 |
| Missing | 0 | 0 | |
| **Somatization (without pain),** mean (SD) | 1.12 (0.72) | 1.57 (0.82) | **0.040** |
| Missing | 0 | 0 | |
| **Somatization (with pain),** mean (SD) | 1.40 (0.67) | 1.64 (0.65) | 0.217 |
| Missing | 0 | 0 | |
| **Somatization (with and without pain),** mean (SD) | 2.52 (1.22) | 3.20 (1.33) | 0.061 |
| Missing | 0 | 0 | |
